# COVID-19 surveillance - a descriptive study on data quality issues

**DOI:** 10.1101/2020.11.03.20225565

**Authors:** Cristina Costa-Santos, Ana Luísa Neves, Ricardo Correia, Paulo Santos, Matilde Monteiro-Soares, Alberto Freitas, Inês Ribeiro-Vaz, Teresa Henriques, Pedro Pereira Rodrigues, Altamiro Costa-Pereira, Ana Margarida Pereira, João Fonseca

## Abstract

**Background:** High-quality data is crucial for guiding decision making and practicing evidence-based healthcare, especially if previous knowledge is lacking. Nevertheless, data quality frailties have been exposed worldwide during the current COVID-19 pandemic. Focusing on a major Portuguese surveillance dataset, our study aims to assess data quality issues and suggest possible solutions.

**Methods:** On April 27^th^ 2020, the Portuguese Directorate-General of Health (DGS) made available a dataset (DGSApril) for researchers, upon request. On August 4^th^, an updated dataset (DGSAugust) was also obtained. The quality of data was assessed through analysis of data completeness and consistency between both datasets.

**Results:** DGSAugust has not followed the data format and variables as DGSApril and a significant number of missing data and inconsistencies were found (e.g. 4,075 cases from the DGSApril were apparently not included in DGSAugust). Several variables also showed a low degree of completeness and/or changed their values from one dataset to another (e.g. the variable ‘underlying conditions’ had more than half of cases showing different information between datasets). There were also significant inconsistencies between the number of cases and deaths due to COVID-19 shown in DGSAugust and by the DGS reports publicly provided daily.

**Conclusions:** The low quality of COVID-19 surveillance datasets limits its usability to inform good decisions and perform useful research. Major improvements in surveillance datasets are therefore urgently needed - e.g. simplification of data entry processes, constant monitoring of data, and increased training and awareness of health care providers - as low data quality may lead to a deficient pandemic control.

## Introduction

The availability of accurate data in an epidemic is crucial to guide public health measures and policies [1]. During pandemics, making epidemiologic data openly available, in real-time, allows researchers with different backgrounds to use diverse analytical methods to build evidence [2,3] in a fast and efficient way. This evidence can then be used to support adequate decision-making which is one of the goals of epidemiological surveillance systems [4]. To ensure that high-quality data are collected and stored, several factors are needed, including robust information systems that promote reliable data collection [5], adequate and clear methods for data collection and integration from different sources, as well as strategic data curation procedures. Epidemiological surveillance systems need to be designed having data quality as a high priority and thus promoting, rather than relying on, users’ efforts to ensure data quality [6]. Only timely, high-quality data can provide valid and useful evidence for decision making and pandemic management. On the contrary, using datasets without carefully examining the metadata and documentation that describes the overall context of data can be harmful [7].

At the moment, producing these high-quality datasets within a pandemic is nearly impossible without a broad collaboration between health authorities, health professionals, and researchers from different fields. The urgency to produce scientific evidence to manage the COVID-19 pandemic contributes to lower quality datasets that may jeopardise the validity of results, generating biased evidence. The potential consequences are suboptimal decision making or even not using data at all to drive decisions. Methodological challenges associated with analysing COVID-19 data during the pandemic, including access to high-quality health data, have been recognized [8] and some data quality concerns were described [7]. Nevertheless, to our knowledge, there is no study performing a structured assessment of data quality issues from the datasets provided by National Surveillance Systems for research purposes during the COVID-19 pandemic. Although this is a worldwide concern, this study will use Portuguese data as a case study.

### The Portuguese systems to input COVID-19 data and the data flows

In early March, the first cases of COVID-19 were diagnosed in Portugal [9]. The Portuguese surveillance system for mandatory reporting of communicable diseases is named SINAVE (National System for Epidemiological Surveillance) and is in the dependence of the Directorate-General of Health (DGS). COVID-19 is included in the list of mandatory communicable diseases to be notified through this system either by medical doctors (through SINAVE MED) or laboratories (SINAVE LAB). A COVID-19 specific platform (Trace COVID-19) was created for the clinical management of COVID-19 patients and contact tracing. However, data from both SINAVE and Trace COVID-19 are not integrated in the electronic health record (EHR). Thus, healthcare professionals need to register similar data, several times, for the same suspect or confirmed case of COVID-19, increasing the burden of healthcare professionals and potentially leading to data entry errors and missing data. The SINAVE notification form includes a high number of variables, with few or no features to help data input. Some examples include 1) within general demographic characteristics, patient occupation is chosen from a drop-down list with hundreds of options and with no free text available; 2) the 15 questions regarding individual symptoms need to be individually filled using a 3-response option drop-down list, even for asymptomatic patients; 3) in the presence of at least one comorbidity, 10 specific questions on comorbidities need to be filled, and 4) there are over 20 questions to characterize clinical findings, disease severity, and use of healthcare resources, including details on hospital isolation. Other examples of the suboptimal design are 5) the inclusion of two questions on autopsy findings among symptoms and clinical signs, although no previous question ascertains if the patient has died; 6) lack of a specific question on disease outcome (only hospital discharge date); 7) lack of validation rules that allow, for example, to have a disease diagnosis prior to birth date or to be discharged before the date of hospital admission and 8) no mandatory data fields, allowing the user to proceed without completing any data. Furthermore, a global assessment of disease severity is included with the options ‘unknown’, ‘severe’, ‘moderate’ and ‘not applicable’ without a readily available definition and without the possibility to classify the disease as mild. This unfriendly system may impair the quality of COVID-19 surveillance data. The problems described have existed for a long time at SINAVE and they are usually solved by personal contact with the health local authorities. However, in the current COVID-19 pandemic scenario, and due to the pressure of the huge number of new cases reported daily, this does not happen at this moment.

Since the beginning of the pandemic, several research groups in Portugal stated their willingness to contribute by producing knowledge and improving data systems and data quality [10]. Researchers requested access to healthcare disaggregated data related to COVID-19, in order to timely produce scientific knowledge to help evidence-based decision-making during the pandemic. On April 27^th^ 2020, the DGS made available a dataset (DGSApril) collected by the SINAVE MED, to be accessed by researchers upon request and after submission of a detailed research proposal and documented approval by an ethical committee [11]. With the DGSApril dataset, DGS also made available the respective metadata [12]. At least 50 research groups received the data and started their dataset analyses.

There are more than one possible data flows from the moment the data are introduced until the dataset is made available to researchers. Figure 1 is an example of the information flow from data introduced by public health professionals until the analysis of data.

**Figure 1:**
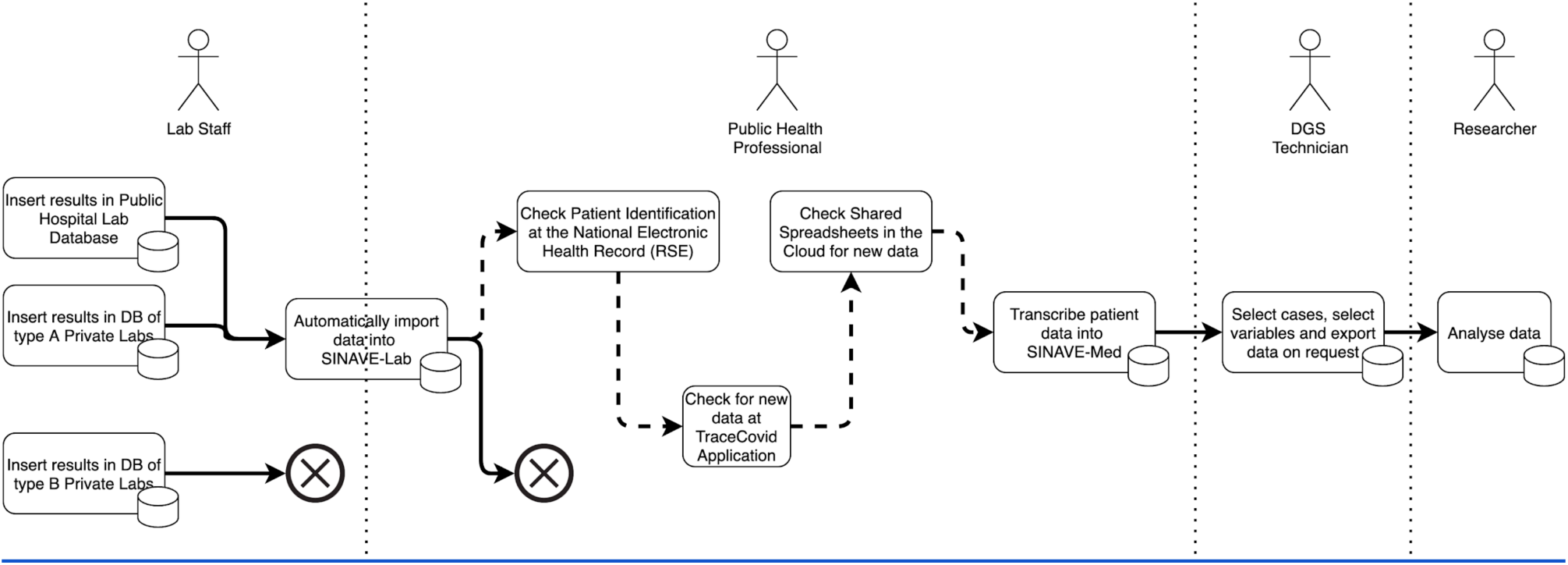
Example of one possible information flow from the moment the data are introduced until the dataset is made available to researchers. The ⊗ symbol means that data are not sent and therefore not present in the research database. The dashed line represents a manual cumbersome process that is many times executed by public health professionals and that is very susceptible to errors.

On August 4^th^ 2020, DGS sent an updated dataset (DGSAugust) to the research groups who had requested the first dataset, including COVID-19 cases already included in the initial dataset plus new cases diagnosed during May and June 2020. This updated database had an inconsistent manifest, including some variables presented in a different format (for example, instead of a variable with the outcome of the patient, the second dataset presented two dates: death and recovery date), or with different definitions (for example, variable age was defined as the age at the time of COVID-19 onset or as age at the time of COVID-19 notification, in the first and second datasets, respectively), which raised concerns regarding their use for valid research and replication of the analysis made using the first version of data.

We aimed to assess data quality issues of COVID-19 surveillance data and suggest solutions to overcome them, using the Portuguese surveillance datasets as an example.

## Methods

The data provided by DGS included all COVID-19 confirmed cases notified through the SINAVE MED and, thus, excluding those only reported by laboratories (SINAVE LAB). The DGSApril dataset was provided on April 27^th^ 2020 and the updated one (DGSAugust) on August 4th 2020. The available variables in both datasets are described in Supplementary File 1.

There was a variable named ‘outcome’, with the information on the outcome of the case, present in DGSApril dataset that was not available in the DGSAugust dataset. On the other hand, there were also some variables (dead, recovery, diagnosis and discharge dates) present in DGSAugust dataset that were not available in the DGSApril dataset.

The quality of the data was assessed through the analysis of data completeness and consistency between the DGSApril and DGSAugust. For data completeness evaluation, missing information was classified as “system missings” when there was no information provided (blank cells) and as “coded as unknown” when the information “unknown” was coded. Considering the consistency, both datasets were compared in order to evaluate if the data quality increased with the update sent four months later. As many data entry errors could be avoided using an optimized information system, the potential data entry errors in DGSAugust were also described.

The number of COVID-19 cases and the number of deaths due to COVID-19 were also compared to the public daily report by Portuguese Directorate-General of Health [13]. We highlight that it is not expected that the daily numbers of cases and deaths reported publicly were coincident to the numbers obtained in the datasets made available to researchers as these datasets included only the COVID-19 cases notified through the SINAVE MED (excluding those only reported by laboratories). However, the calculation of this difference is important to estimate the potential bias that data of these (DGSApril and DGSAugust) datasets, provided by DGS to researchers, may have. This comparison is only possible in the DGSAugust dataset as in the DGSApril dataset the variable date of diagnosis was not available.

## Results

### Cases included and omitted

From the 20,293 COVID-19 cases included in the DGSApril dataset, only 80% (n=16,218) had the same unique case identifier in the DGSAugust dataset. There were 4,075 cases in the DGSApril dataset that were not included in the DGSAugust dataset or, alternatively, had changed the unique case identifier. The DGSAugust dataset provided a total of 38,545 COVID-19 cases, including 22,327 that were not available in DGSApril dataset: 5,713 diagnosed until April 27^st^ but that presumably were not included in the DGSApril dataset, 16,609 diagnosed after the period included in the DGSApril dataset, and 5 cases with missing information on diagnosis date (Figure 2).

**Figure 2:**
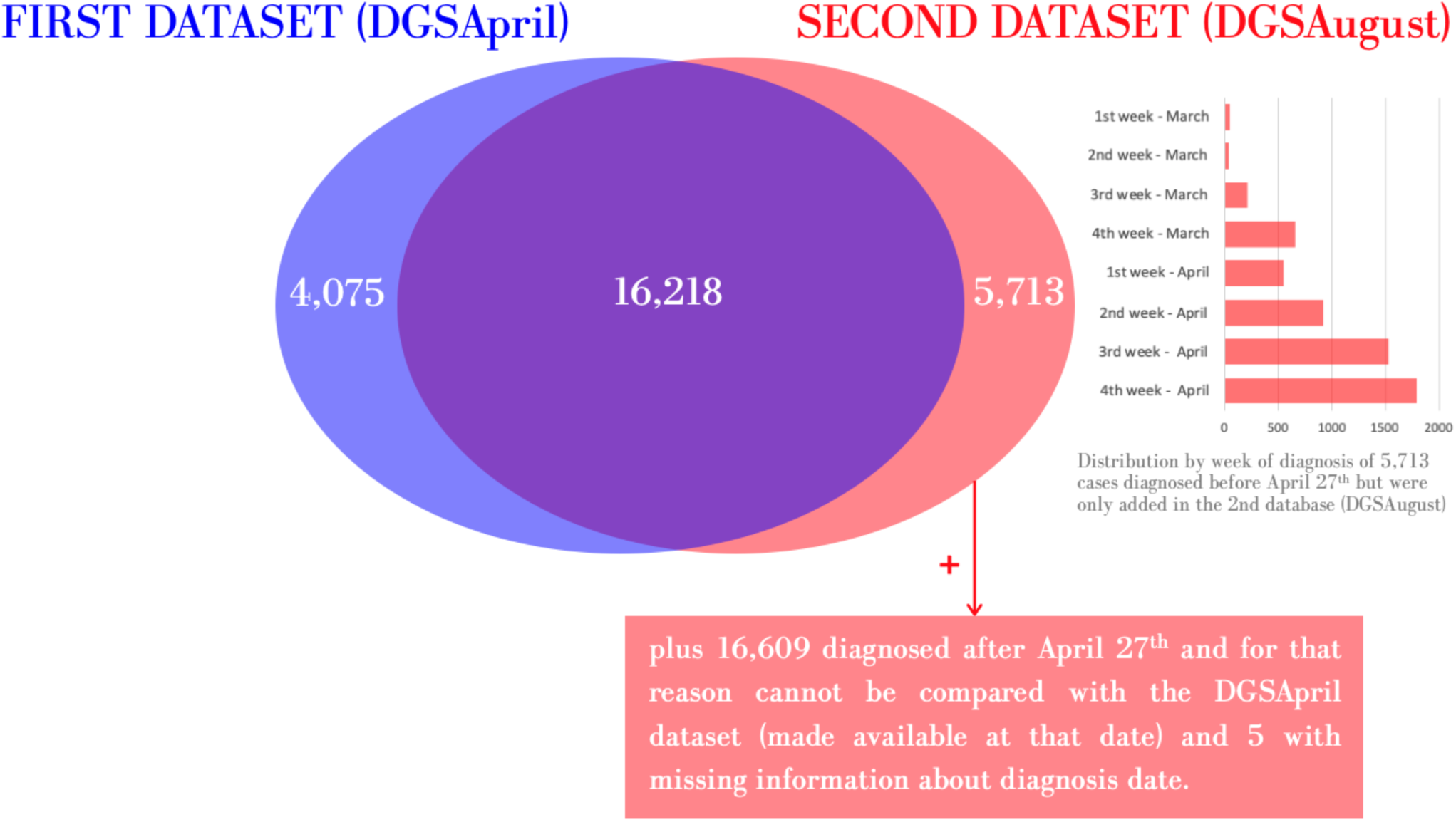
Number of unique case identifiers presented in the datasets of COVID-19 cases diagnosed since the start of the pandemic until April 27^th^ (date when the first database was made available) and after April 27^th^

Considering the 5,713 cases made available only in the DGSAugust and diagnosed before April 27th that, presumably, were not included in the DGSApril dataset, the majority (58%) were diagnosed in the two weeks immediately prior to the April 27th (the date on which this database was made available). However, 42% were diagnosed more than two weeks before the DGSApril dataset was made available (Figure 2).

### Data completeness of both datasets

Several variables showed a low degree of completeness. For example, two variables (“date of first positive laboratory result” and “case required care in an intensive care unit”) had more than 90% of cases with missing information in DGSApril dataset - coded as unknown or system missing. In the DGSAugust dataset, the variable ‘case required care in an intensive care unit’ reduced the proportion of incomplete information to 26% of system missings and no cases were coded as unknown. However, the variable ‘Date of first positive laboratory result’ still had 90% system missings in the DGSAugust dataset. Table 1 provides detailed information about missing information for each available variable.

**Table 1:** Data completeness (number and percentage of missing information) of each variable available in the DGSApril and DGSAugust datasets with COVID-19 cases provided by DGS.

|  | DGSApril<br>n=20293 |  | DGSAugust<br>n=38545 |  |
| --- | --- | --- | --- | --- |
|  | System<br>missing<br>n (%) | Coded as<br>unknown<br>n (%) | System<br>missing<br>n (%) | Coded as<br>unknown<br>n (%) |
| Unique case identifier (Recordid) | 0 | 0 | 0 | 0 |
| RecordID of the linked cases | a) | a) | a) | a) |
| Age | 0 | 0 | 0 | 0 |
| Probable place of infection | 0 | 0 | 0 | 0 |
| Gender | 0 | 0 | 0 | 0 |
| Hospitalization | 0 | 1623 (8) | 3 (0) | 3425 (9) |
| Outcome | 0 | 23 (0) | b) | b) |
| Patient has underlying condition | 0 | 2 (0) | 15407 (40) | 2495 (6) |
| Date of first positive laboratory result | 19268 (95) | 0 | 34667 (90) | 0 |
| Date of diagnosis | c) | c) | 7 (0) | 0 |
| Date of disease onset | 4815 (24) | 0 | 15045 (39) | 0 |
| Date of death | c) | c) | 37390 (97) | 0 |
| Date of recovery | c) | c) | 21499 (56) | 0 |
| Only hospitalized cases | n=2973 |  | n=4327 |  |
| Date of hospitalization | 386 (13) | 0 | 860 (20) | 0 |
| Case required care in an intensive care unit | 0 | 2712 (91) | 1122 (26) | 0 |
| Level of respiratory support given to patient | 0 | 1573 (53) | 1364 (31) | 172 (4) |
| Date of hospital discharge | c) | c) | 3975 (92) | 0 |

a) variable neither available in DGSApril dataset nor in DGSAugust dataset but described in the metadata file provided by DGS
b) variable available in DGSApril dataset and described in the metadata file provided by DGS but not provided in DGSAugust dataset
c) variable neither available in DGSApril dataset nor described in the metadata file provided by DGS but provided in DGSAugust dataset

### Data consistency between DGSApril and DGSAugust datasets

The consistency of the information for cases identified with the same unique case identifier in both datasets (n=16,218) was further evaluated (Figure 1). Table 2 presents the number and percentage of cases with different information, for each variable.

**Table 2:**
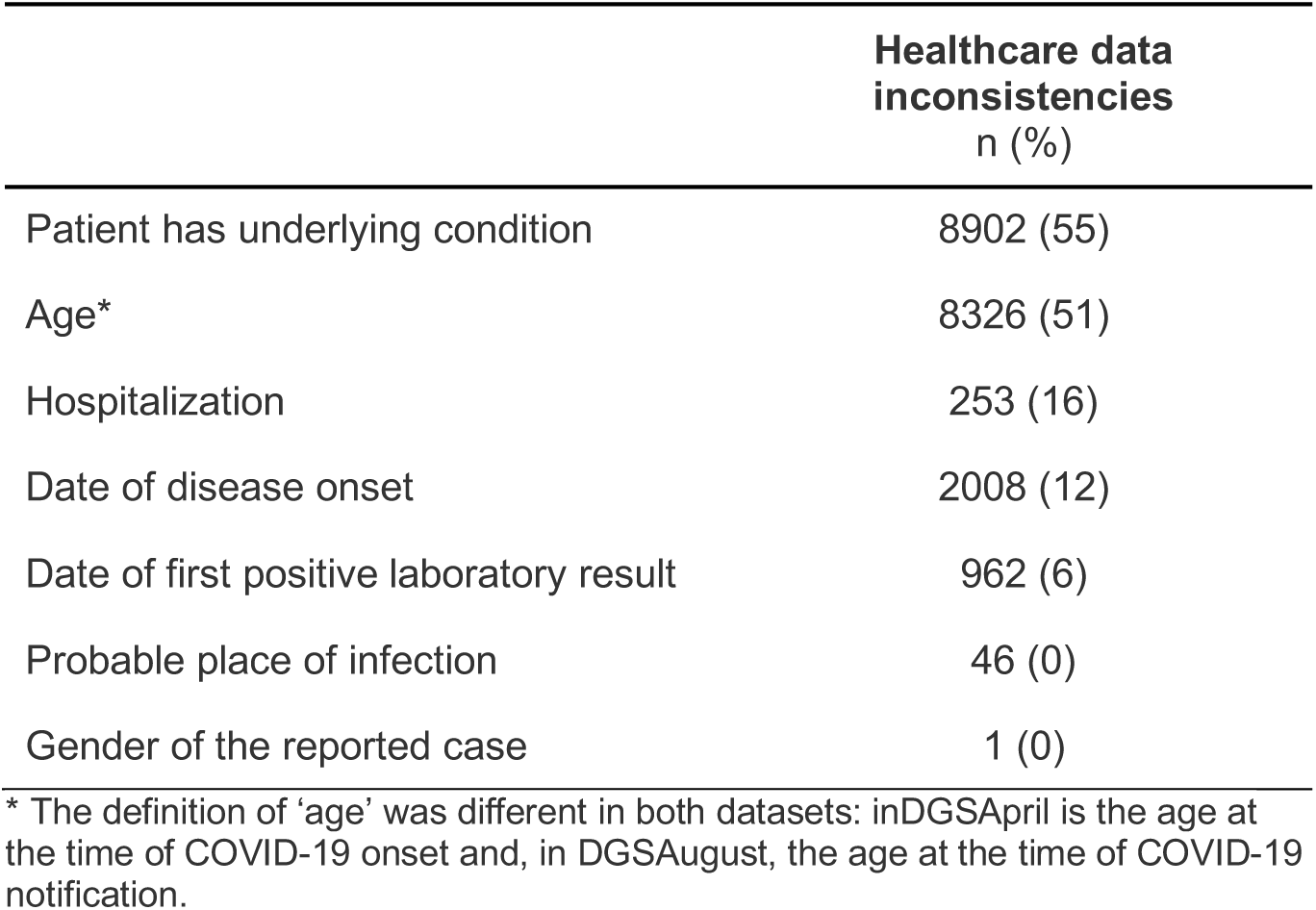
Number and percentage of COVID-19 cases presented in both datasets (n=16218) with information that did not match for each variable.

|  | **Healthcare data inconsistencies**<br>n (%) |
| --- | --- |
| Patient has underlying condition | 8902 (55) |
| Age* | 8326 (51) |
| Hospitalization | 253 (16) |
| Date of disease onset | 2008 (12) |
| Date of first positive laboratory result | 962 (6) |
| Probable place of infection | 46 (0) |
| Gender of the reported case | 1 (0) |

The variable ‘underlying conditions’ was the one showing a higher percentage of inconsistencies between both datasets, with more than half of cases showing different information when comparing the information from both datasets (Table 2). Most of the inconsistencies were due to the cases recorded as ‘no underlying conditions’ in the DGSApril dataset and corrected to ‘Unknown if the case has underlying conditions’ or ‘missing’ in the updated dataset (DGSAugust) (42%, n=6851). There were 1,952 cases (12%) recorded as ‘no underlying conditions’ in the first dataset and corrected to ‘yes - underlying conditions’ in the second one. There were also 99 (1%) cases with underlying conditions in the first dataset corrected to ‘no underlying conditions’ in the second one. The variable ‘age’ also had more than half of cases showing different information when comparing the information from both datasets (Table 4). The difference in all cases with different information, except one, was 1 year old. The definition of ‘age’ was different in both datasets: in DGSApril is the age at the time of COVID-19 onset and, in DGSAugust, the age at the time of COVID-19 notification.

**Table 3:**
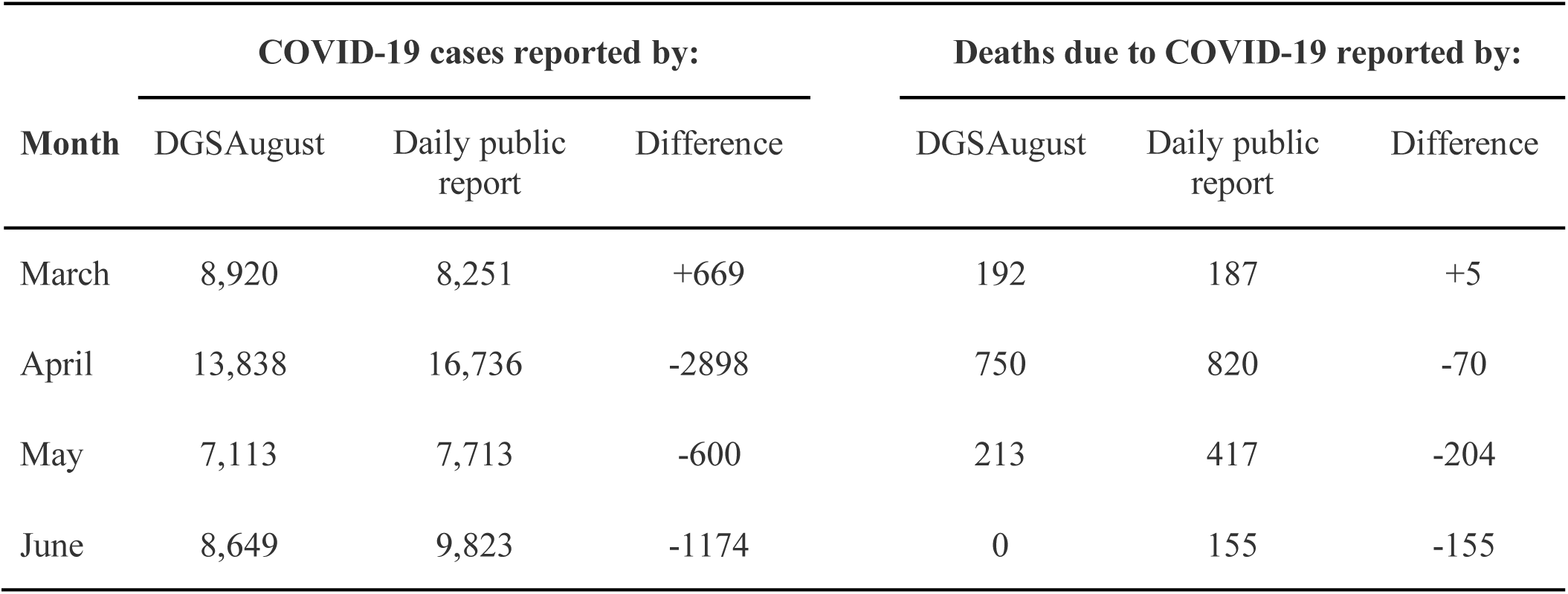
Number of COVID-19 cases and deaths due to COVID-19 reported by DGSAugust dataset and by the daily public report.

**Table 4:**
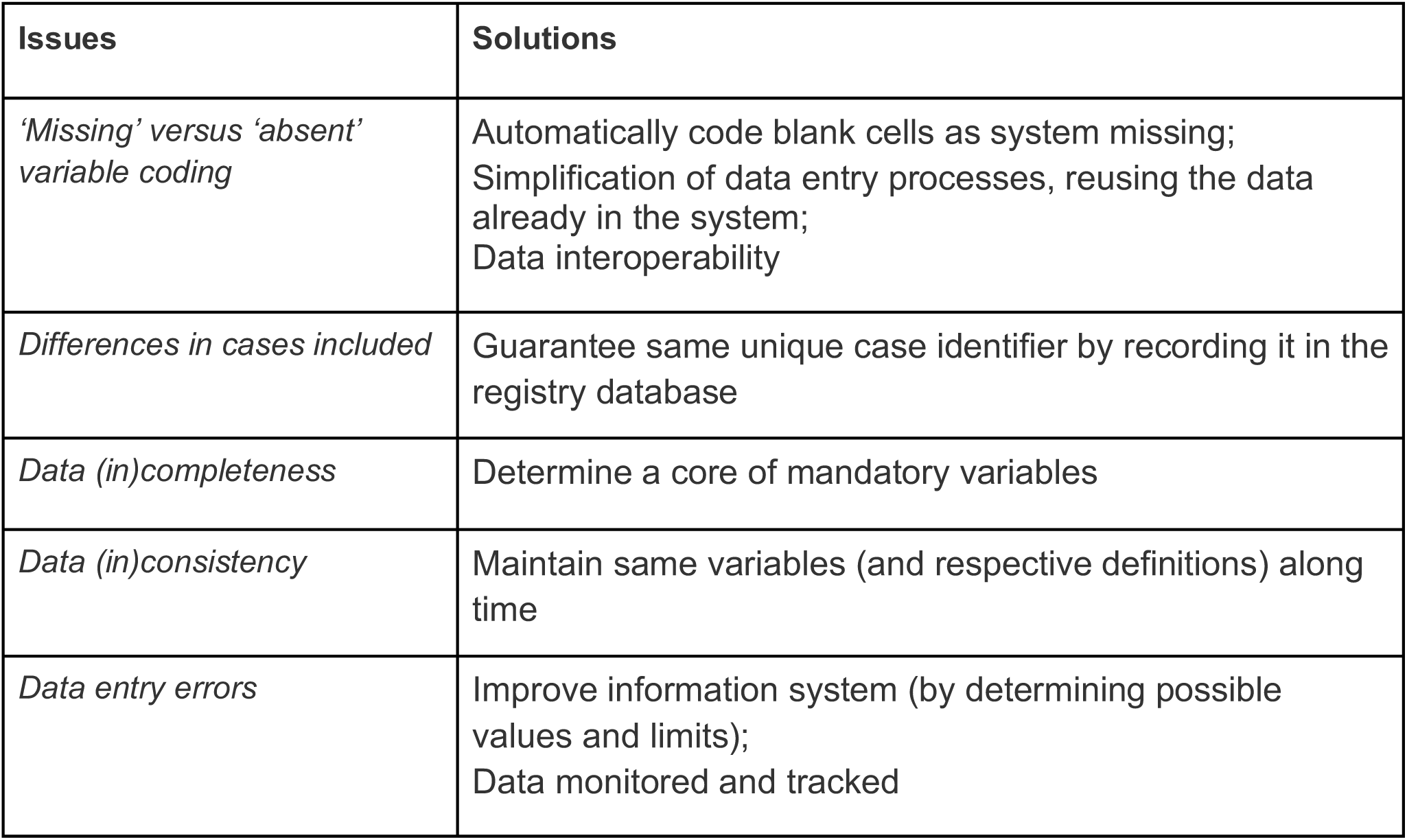
Most frequent data quality issues and possible solutions

| Issues | Solutions |
| --- | --- |
| *'Missing' versus 'absent' variable coding* | Automatically code blank cells as system missing;<br>Simplification of data entry processes, reusing the data already in the system;<br>Data interoperability |
| *Differences in cases included* | Guarantee same unique case identifier by recording it in the registry database |
| *Data (in)completeness* | Determine a core of mandatory variables |
| *Data (in)consistency* | Maintain same variables (and respective definitions) along time |
| *Data entry errors* | Improve information system (by determining possible values and limits);<br>Data monitored and tracked |

The variable ‘hospitalization’ had 16% of cases (n=253) with unmatched information (Table 2). One hundred and twenty-five cases were recorded as ‘Unknown if the case was hospitalized’ in the DGSApril dataset and corrected to ‘No hospitalization’ in the DGSAugust. Sixty-two cases were recorded as “No hospitalization” and corrected to ‘Hospitalized’ or ‘Unknown information’ in DGSApril and DGSAugust datasets, respectively. Fifty-five cases were recorded as hospitalized patients and corrected to ‘No hospitalization’ or ‘Unknown information’ in DGSApril and DGSAugust datasets, respectively. Only 11 cases changed from ‘Unknown if the case was hospitalized’ to ‘Hospitalization’. The variable ‘date of disease onset’ had 12% of cases (n=2008) with unmatched information (Table 2). In 1,445 cases, information about the date of disease onset was provided only in DGSApril and 563 cases had dates in both datasets but the dates did not match.

The variable ‘date of the first positive laboratory result’ did not match in both datasets in 6% of the cases (n=962). In 5 cases there was a date available in both datasets but the dates did not match, in 74 cases the date was available only in the DGSApril dataset, and in 883 cases the date was available only in the DGSAugust dataset.

The variable patient outcome (variable ‘outcome’) was not present in the DGSAugust dataset which instead presents the variables ‘date of recovery’ and ‘date of death’ (not presented in DGSApril) (Table 1). In the DGSApril dataset, there were 1,134 cases coded as ‘Alive, recovered and cured’, but only 83% of those (n=947) had recovery date in the updated dataset (DGSAugust), which may be due to the lack of information on a specific date, despite knowing that the case result is alive, recovered and cured. In fact, 177 patients recorded as ‘alive, recovered and cured’ in the DGSApril, did not have any date in the DGSAugust dataset. However, 10 patients recorded as ‘alive, recovered and cured’ in the DGSApril had a date of death in the DGSAugust dataset. Seven of these were dates of death before April 19, which is incongruent. Among the 455 cases coded as ‘Died because of COVID-19’ in the DGSApril dataset, 7 (2%) did not have a date of death in the second dataset.

### Data entry errors in the updated dataset (DGSAugust)

The age of one patient is probably wrong (134 years old). There were 3 male patients and an older woman (97 years old) registered as pregnant.

There was a wrong diagnosis date (50-05-2020) and 19 patients had registered dates of diagnosis before the first official case of COVID-19 was diagnosed in Portugal. There were also two patients with a negative length of stay in hospital.

The variable ‘recovery date’ had only three values even though it refers to a 120 days períod - for 6,772 patients the date of recovery was recorded as ‘April 3’, for 1,032 patients ‘May 25’ and for 242 the date of recovery recorded was ‘May 26’.

### Number of COVID-19 cases provided by DGSAugust dataset and by daily public report

Table 3 shows the number of COVID-19 cases reported by DGSAugust dataset and by the daily public report. The DGSAugust dataset included 38520 COVID-19 cases diagnosed between March and June, less 4,003 cases (9%) than th*e* daily public report provided by Portuguese Directorate-General of Health. However, when looking at data from March, the DGSAugust dataset reported more 669 cases (8%) than th*e* daily public report. In April, May and June the DGS dataset reported less 17%, 8% and 12% of cases than the public report provided, respectively.

### Number of deaths due to COVID-19 cases provided by DGSAugust dataset and by daily public report

Table 3 shows the number of deaths due to COVID-19 reported by DGSAugust dataset and by the daily public report. The DGSAugust dataset reported 1,155 deaths due to COVID-19 until the end of June, less 424 cases (27%) than th*e* daily public report provided by the Portuguese Directorate-General of Health. However, in March the DGSAugust dataset reported more 5 deaths due to COVID-19 (3%) than th*e* daily public report. In April, May and June the DGS dataset reported less 8%, 49% and 100% of cases than the public report provided, respectively.

## Discussion

The production of scientific evidence to help manage the COVID-19 pandemic is an urgency worldwide. However, if the quality of datasets is low the evidence produced may be inaccurate and, therefore, have limited applicability. This problem may be particularly critical when low-quality datasets provided by official organisations lead to the replication of biased conclusions in different studies.

The problem of using datasets with suboptimal quality for research purposes during the COVID-19 pandemic probably occurs in a large number of countries. This study, using the Portuguese surveillance data, reports a high number of inconsistencies and incompleteness of data that may interfere with scientific conclusions. To date, we could identify three scientific papers reporting analysis of these data [14,15,16] that may have been affected by the low quality of the datasets [17]. Table 4 presents data quality issues identified in the provided datasets and possible solutions.

The issue of ‘missing’ versus ‘absent’ variable coding seems to be present in the findings of the paper by Nogueira et al. [14] The reduction of the risk of death in relation with comorbidities observed on the analysis of first dataset is underestimated if we assume that the updated dataset is the correct. [17]. If this analysis had included the 6,851 cases as missing values, the results and conclusions could be indeed different. In fact, these cases were registered as having no underlying conditions in the first dataset but corrected in the second dataset to ‘Unknown if the case has underlying conditions’ or system missing. This problem might be due to the way these data were collected and/or were recorded in the database sent to the researchers. In the form used to collect COVID-19 surveillance data, comorbidities are recorded one by one after a general question assessing the presence of any comorbidity and the field is not mandatory. From a clinical point of view, it might be enough to register only positive data perceived as relevant (e.g. the presence of a specific diagnosis, but not its absence), especially in a high-burden context as the ongoing pandemic. In the context of clinical research, however, the lack of registered comorbidity data cannot be interpreted as the absence of comorbidities. A similar bias can be found in the other two studies reporting analysis of DGSApril dataset [15,16].

Another data quality issue is related with the differences in cases included. In fact, only 80% of cases included in the DGSApril dataset had the same unique case identifier in the DGSAugust dataset and only 74% of cases diagnosed until April 27^th^ included in DGSAugust had the same unique case identifier in the DGSApril. Alternatively, the unique case identifier had been changed. We do not know if the unique identifier is generated in each data download or if it is recorded in the database. This last option will be the safest. Moreover, until June 19, it was not mandatory to fill in the national health service user number in order to have a standard unique patient identifier. That may have led to not identifying duplicate SINAVE MED entries for the same patient and increased the difficulty in adequately merging data from SINAVE LAB, SINAVE MED and other data sources.

The high percentage of incomplete data in several variables may also produce biases whose dimensions and directions are not possible to estimate. In fact, as our results showed, half of the variables available in the DGSAugust dataset had more than one-third of missing information. Furthermore, that dataset was already incomplete since it only provides COVID-19 cases from the medical component of SINAVE totalizing 90% of the cases reported by health authorities until the end of June 2020 [13]. It is unclear, however, why the updated version of the dataset in March reported more 669 COVID-19 cases and more 5 deaths than the public report (which would be expected to be more complete). Moreover, there were no reported dates of deaths in June in DGSAugust dataset, despite the 155 deaths reported in the public report during this month.

The consistency of variables in different updates of datasets is also an important quality issue. In fact, our results shows that the variable ‘age’ was calculated differently in the two datasets: in the DGSApril dataset it was the age at the time of COVID-19 onset and in the DGSAugust dataset it was age at the time of COVID-19 notification. Despite this change in definition, the difference of one year in half of the cases does not seem to be completely justified only by this fact, since the two dates should be relatively close. Still related to this problem of inconsistent information and variables, we realised that some information may have been lost in the second dataset sent (DGSAugust). In fact, the outcome of the COVID-19 case is not presented in the second dataset. DGSAugust dataset only presents the recovery and death dates. It would be possible to reconstruct in the second dataset some of the information on the outcome variable presented in the first one. However, it would only be possible to directly recode those with ‘Date of recovery’ as ‘Alive recovered cured’; all other categories (‘Died of COVID-19’; ‘Died of other cause’; ‘Cause of death unknown’; ‘Still on medical treatment’) are impossible to obtain from the dates of recovery or death. In fact, using only the variable ‘date of death’, it is not possible to determine if the patient died because of COVID-19, died of another cause, or if the cause of death is unknown as in the DGSApril dataset. Moreover, 17% of cases coded as ‘Alive, recovered and cured’ in the first dataset did not have the variable ‘Date of recovery’ filled in the updated one. While the recovery date (when available) can be used as a proxy of the patient outcome, if this date is unknow in spite of a known recovery we miss the whole outcome information.

In fact, in the DGSAugust dataset it is assumed that the missing information about the recovery date implies that the case had not recovered yet. Also, the ‘recovery date’ had only three dates even though it refers to a four-month period.

All the described errors, inconsistencies, data incompleteness, changes in the variables’ definitions and format may lead to unreproducible methods and analyses. While important to start working in data analysis as fast as possible in the early beginning of a pandemic, it is also crucial that the models and analysis developed with the first data are validated *a posteriori* and confirmed with the updated data. It is thus fundamental that the subsequent datasets follow the same metadata and preferably are more complete and with less inconsistencies and errors.

Quality of healthcare data can be improved through several strategies. First, data entry processes must be simplified, avoiding duplications and reusing the data already in the system, since the need to input the same information in different systems is time-consuming, frustrating for the user, and can negatively impact both data completeness and accuracy. Data interoperability can also be a powerful approach to minimise the number of interactions with the system [18]. Second, data needs to be constantly monitored and tracked [19]: organisations must develop processes to evaluate data patterns, and establish report systems based on data quality metrics. Even before data curation, simple validation procedures and rules in information systems can help detecting and preventing many errors (i.e. male patients classified as “pregnant”, or a patient aged 134 years old) and inconsistencies, and improve data completeness.

Finally, we need to establish the value proposition for both creators and observers [20]. This includes ensuring that healthcare providers understand the importance of data, receive feedback about their analysis and how it may improve both the assistance to the patient and the whole organization, and have received adequate training for better performance.

The adoption of these strategies should pave the way to high-quality, accurate healthcare datasets that can generate accurate knowledge to timely inform health policies, and the readaptation of health care systems to new challenges.

Our study has some limitations. We asked DGS for clarification on some data issues and are still waiting to receive complete answers that might clarify some of these aspects. Therefore, the analysis of the Portuguese surveillance data quality was done exclusively with the analysis of the databases provided by DGS to researchers and with our external knowledge about how the information flows from the moment the data are introduced by health professionals until the dataset can be used for data analysis. Another limitation is the fact that we only studied the quality issues of COVID-19 data from one country, Portugal. However our results seem to be in line with the findings of Ashofteh and colleagues [7] who analysed and compared the quality of official datasets available for COVID-19, including data from the Chinese Center for Disease Control and Prevention, the World Health Organization,and the European Centre for Disease Prevention and Control. In fact, they also found noticeable and increasing measurement errors in the three datasets as the pandemic outbreak expanded and more countries contributed data for the official repositories.

## Conclusion

We describe some important quality issues of the Portuguese COVID-19 surveillance datasets, which may jeopardize the validity of some analysis, with possible serious implications in a context as a pandemic.

The availability of official data by National Health Authorities to researchers is an enormous asset, allowing data analysis, modelling and prediction, that may support better decisions for the patient and the community as a whole. However, to fully embrace this potential, it is crucial that these data are accurate and reliable.

It urges to define and implement major improvements in the processes and systems of surveillance datasets: simplification of data entry processes, constant monitoring of data, raise awareness of health care providers for the importance of good data and providing them adequate training.

Data curation processes, capitalising on effective and multidisciplinary collaborations between healthcare providers and data analysts, play a critical role to ensure minimum quality standards. Once these processes are fully optimised, the reliability of results and the quality of the scientific evidence produced can be greatly improved.

## Data Availability

Data used in this work was made available by the Portuguese Directorate-General of Health, under the scope of article 39th of the decree law 2-B/2020, from April the 2nd and is available from request.

## Acknowledgments

Data used in this work was made available by the Portuguese Directorate-General of Health (DGS), under the scope of article 39th of the decree law 2-B/2020, from April the 2nd.

