## Supplementary File 1 for "COVID-19 surveillance - a descriptive study on data quality issues"

Table A: Description of variables available in DGSApril dataset.

| **FIRST DATASET** | |
|---|---|
| **Variable name** | **Variable description (type of variable)** |
| Recordid | Unique case identifier (TEXT) |
| Age | Age of patient in years as reported in the national system at the time of disease onset.<br>(NUMERICAL) |
| AgeDay | Age of patient in days as reported in the national system for cases < 1 month of age at the time of disease onset (NUMERICAL) |
| AgeMonth | Age of patient in months as reported in the national system for cases < 2 years of age at the time of disease onset (NUMERICAL) |
| DateOfFirstPositiveLabResult | Date when first positive laboratory result become available (DATE) |
| DateOfHospitalisation | Date of Hospitalisation (DATE) |
| DateOfOnset | Date of onset of disease. Not applicable in asymptomatic cases. If not applicable, please use 'Unk' (DATE) |
| Gender | Gender of the reported case (CODED VARIABLE)<br>*Codes: F=Female; M=Male; Unk=Unknown* |
| Hospitalisation | Admission to hospital (CODED VARIABLE)<br>*Codes: N= No; Unk=Unknown; Y=Yes* |
| IntensiveCare | Case required care in an intensive care unit (CODED VARIABLE)<br>*Codes: N= No; Unk=Unknown; Y=Yes* |
| Outcome* | Information on the outcome of the case (CODED VARIABLE)<br>*Codes: ALIVE= Alive, Recover, cured; DIEDNCOV = Died because of new coronavirus; DIEDOTHER = Died because of other cause; DIEDUNK = Cause of death unknown; STILLTREATMENT = Still on medical treatment (not recovered); UNK = Unknown outcome* |
| PlaceOfInfection | The probable place of infection should be provided at the NUTS 3 level. If out of the country, it will be used country codes (CODED VARIABLE)<br>*Codes: NUTS III codes* |
| Precondition | Patient's underlying condition or conditions (CODED VARIABLE)<br>*Codes: CANC=Cancer, malignancy; CARDIACDIS=Cardiac disorder, including hypertension; DIAB=Diabetes; HIV=HIV/other immune deficiency, KIDNEY=Kidneyrelated condition, renal disease; LIVER=Liver-related condition, liver disease; LUNG=Chronic lung disease; NEUROMUS=Neuromuscular disorder, chronic neurological;NONE=None; O=Other signs, please specify; PREG=Pregnancy, trimester is unknown; PREG1=Pregnancy, 1st 3 trim, the 1st trim is from week 1 to the end of week 12; PREG2=Pregnancy, 2nd trim, the 2nd trim is from week 13 to the end of week 26; PREG3=Pregnancy, 3rd trim, the 3rd trim is from week 27 to the end of the pregnancy; PREGPOST=Postpartum (<6 weeks); UNK=Unknown precondition* |
| PreconditionOther | Details of underlying conditions, if Precondition is coded as ‘other’, but is known (TEXT) |
| RespSupport | Level of respiratory support given to patient (CODED VARIABLE)<br>*Codes: N=No; O=Other, please specify; OXYGEN=Oxygen therapy; UNK=Unknown; VENT=Ventilator including non-invasive pos pressure vent* |

* not available in DGSAugust dataset.

Table B: Description of variables available in DGSAugust dataset.

| **FIRST DATASET** | |
|---|---|
| **Variable name** | **Description** |
| Recordid | Unique case identifier (NUMERICAL) |
| Age | Age of patient in years, days or months as reported in the national system at the time of notification (TEXT) number with the units (years/months/days) |
| DateOfFirstPositiveLabResult | Date when first positive laboratory result become available (DATE) |
| DateOfDead* | Dead date (DATE) |
| DateOfRecovery* | Recovery date (DATE) |
| DateOfHospitalisation | Date of Hospitalisation (DATE) |
| DateOfOnset | Date of onset of disease (DATE) |
| DateOfDiagnosis* | Date of Diagnosis (DATE) |
| DateOfDischarge * | Discharge date (DATE) |
| Gender | Gender of the reported case (CODED VARIABLE)<br>*Codes: F=Female; M=Male* |
| Hospitalisation | Admission to hospital (CODED VARIABLE)<br>*Codes: Unk=Unknown; Y=Yes; N=No;* |
| IntensiveCare | Case required care in an intensive care unit (CODED VARIABLE)<br>*Codes: 1=Yes; 0=No;* |
| PlaceOfInfection | The probable place of infection should be provided at the NUTS 3 level. If out of the country, it will be used country codes (CODED VARIABLE)<br>*Codes: NUTS III codes* |
| Precondition | Patient have underlying condition or conditions (CODED VARIABLE)<br>*Codes: Unk=Unknown; Y=Yes; N=No;* |
| Precondition: cancer | Patient's underlying condition: Cancer, malignancy (CODED VARIABLE)<br>*Codes: Unk=Unknown; Y=Yes; N=No* |
| Precondition: Diabetes | Patient's underlying condition: Diabetes (CODED VARIABLE)<br>*Codes: Unk=Unknown; Y=Yes; N=No* |
| Precondition: HIV | Patient's underlying condition: HIV (CODED VARIABLE)<br>*Codes: Unk=Unknown; Y=Yes; N=No* |
| Precondition: Neuromuscular | Patient's underlying condition: Neuromuscular disorder, chronic neurological (CODED VARIABLE)<br>*Codes: Unk=Unknown; Y=Yes; N=No* |
| Precondition: Asma | Patient's underlying condition: Asma (CODED VARIABLE)<br>*Codes: Unk=Unknown; Y=Yes; N=No* |
| Precondition: Lung | Patient's underlying condition: Chronic lung disease (CODED VARIABLE)<br>*Codes: Unk=Unknown; Y=Yes; N=No* |
| Precondition: Liver | Patient's underlying condition: Liver disease (CODED VARIABLE)<br>*Codes: Unk=Unknown; Y=Yes; N=No* |
| Precondition: Hematologic | Patient's underlying condition: Hematologic disease (CODED VARIABLE)<br>*Codes: Unk=Unknown; Y=Yes; N=No* |

| | |
|---|---|
| Precondition: Kidney | Patient's underlying condition: Kidney disease (CODED VARIABLE)<br>*Codes: Unk=Unknown; Y=Yes; N=No* |
| Precondition: Neurologic | Patient's underlying condition: Chronic neurological deficiency (CODED VARIABLE)<br>*Codes: Unk=Unknown; Y=Yes; N=No* |
| Precondition: Other | Other patient's underlying condition or conditions (TEXT) |
| Precondition: Pregnancy | Pregnancy (CODED VARIABLE)<br>*Codes: Unk=Unknown; Y=Yes; N=No* |
| Precondition: Gestational age | Gestational age (CODED VARIABLE)<br>*Codes: first=1 to 12 weeks; second=13 to 26 weeks; third=27 or more weeks; Unk=Unknown; Post_childbirth: delivery less than 6 weeks ago* |
| RespSupport | The patient need respiratory support (CODED VARIABLE)<br>*Codes: N=No; Y=Yes* |
| RespSupportWhich | Which respiratory support (TEXT) |

* not available in DGSApril dataset.
